# Systematic searching in Ovid Embase: understanding the MEDLINE document collection and identifying conference records

**DOI:** 10.1101/2022.04.23.22274201

**Authors:** Paul Levay

## Abstract

Ovid Embase is an important database in systematic searches for health and medical topics. This paper shows how the document collections on Ovid Embase can be used to limit searches and isolate the unique content from MEDLINE that is available on Embase. This is used to test a method for identifying conference records from Embase. The recommended method found 5,108,945 conference records on Embase on Ovid March 1 2022, about 14% of the database. Search strategies are provided for the scenarios that are most likely to be encountered in systematic searches.

## Introduction

### Background

Embase is an important database in systematic searches for health and medical topics.^1^ Embase is recommended as a source when searching for Cochrane Reviews of Interventions,^2^ Campbell Systematic Reviews,^3^ JBI systematic reviews and evidence syntheses,^4^ and health technology assessments on clinical effectiveness.^5^ Embase, produced by Elsevier, is available through several platforms, such as EBSCOhost, Embase.com, ProQuest and Ovid. This paper refers specifically to the Ovid version of the database, as the search functions and other features are likely to differ on the other platforms.^6^

Embase has been found to be one of the key options for identifying conference records, even though it may still be worth manually checking journal supplements and other sources.^7^ It is useful to know how to identify conference records comprehensively, whether the intention is to include or exclude them from a set of search results.

### Identifying conference records

The systematic searches to support systematic reviews, meta-analyses and other types of evidence synthesis need to identify as much of the relevant evidence as possible.^2^ Identifying conference records can be an important way of minimizing publication bias to ensure evidence syntheses are based on comprehensive search results. Publication bias means that research with statistically significant or clinically favorable results is more likely to be published than research with non-significant or unfavorable results.^8^

Conference records can be time consuming to identify and process, which means they may not be used in all evidence syntheses.^9^ Review teams need to decide how to treat data that is exclusively available from conference records. Identifying conference records separately can facilitate this process. A comprehensive search for conference records may be more helpful in systematic reviews of interventions with small effect sizes or conflicting evidence.^9^ Running separate searches enables the reviewers to conduct a sensitivity analysis so that the meta-analysis can be run with and without the unique data from conference records.^9^

Scoping reviews or mapping reviews that aim to understand the evidence base may also benefit from separate searches, as these will show the availability of full studies compared to conference records. Conference records may be useful, even if they are not included in their own right, for example if they identify key authors and principal investigators, who are then contacted to request full data.^10^

Administering search results might be easier if conference records are treated separately. It can be difficult to remove duplicates from search results, as conference records sometimes have the same title as later papers. It could also be beneficial to treat conference records in isolation when machine learning technology is being used to prioritize screening. Conference records may not have an abstract (or the abstract may actually be the full text) and this affects the effectiveness of machine learning technology.^11^

### The components of Embase

In order to search Embase accurately, it is important to understand how the database is constructed. Elsevier provide bibliographic information, abstracts and indexing using the Emtree thesaurus for around 8,500 journals.^12^

Embase is an important source of materials presented at conferences, including abstracts of presentations and the text of posters. The Ovid guide to Embase suggests that Elsevier process around 350,000 conference records a year.^12^ This number has, of course, been lower in recent years because of the COVID-19 pandemic. Embase has included this content since 2009 and it is often found in supplements to journals, in the form of an abstract. The conference abstract often constitutes the whole record, as there in no further full text.^13^ Automated processes are used to index the conference records.^13^

Embase also contains a supplement of content added to MEDLINE since 2010, which provides records from a further 2,400 journals.^12^ Elsevier does not index the unique MEDLINE content separately but it does use an automated process to map the subject headings from MeSH to Emtree.^13^ This means that the unique MEDLINE content is not indexed with Emtree-specific terms that do not occur in MeSH.^13(p12)^

In 2022, Elsevier started adding records of preprints to Embase. Preprints are preliminary manuscripts that have not undergone formal peer review. The content includes papers added to the preprint repositories *medRxiv* and *bioRxiv* since 28 December 2021. Elsevier index this preprint content, as well as articles-in-press, using automated processes when adding it to Embase.^14^

Embase records are added to four document collections: *Conference Abstracts, Embase, MEDLINE* and *Preprints (unpublished, non-peer reviewed)*. These document collections are accessed by using the *Records From* limit in Ovid Embase. This limit applies the definitions in Table 1 to the search results. Librarians and other database administrators can configure the *Records From* limits display at an institutional level to ensure their users have the optimal search experience with this feature.

**Table 1.** Definitions used in the *Records From* limit in Ovid Embase.

| Limit | Definition |
| --- | --- |
| *Records From* Conference Abstracts | Conference Abstract (records identified as uniquely Conference Abstract and are Elsevier copyrighted) |
| *Records From* Embase | Embase (records indexed by Embase and are Elsevier copyrighted; these records may also have been indexed separately by MEDLINE) |
| *Records From* MEDLINE | MEDLINE (records identified as uniquely MEDLINE and have not been indexed by Elsevier) |
| *Records from* Preprints (unpublished, non-peer reviewed) | Preprints (unpublished, non-peer reviewed) (records identified as uniquely Preprint and are Elsevier copyrighted) |

The inclusion of MEDLINE content may influence how Embase is searched most effectively. A survey published in 2018 suggests that, although MEDLINE content could be accessed through Embase, searchers were reluctant to use the databases together.^15^ It follows that searchers who want to use the databases separately may prefer to remove the unique MEDLINE content from their Embase search results, particularly as a way to manage the number of duplicate records to be processed.^16^

Premji and Ganshorn tested the reliability of using *Records From* Embase and found it could be appropriate to use it in systematic reviews.^16^ Premji and Ganshorn acknowledge that this method must be applied carefully when the purpose is to either exclude confererence records or to identify them separately.^16(p10)^

This paper provides further information on using the *Records From* Embase limit, then demonstrates a method for comprehensively identifying conference records.

## Methods

### Aims and objectives

The aim was to identify a method of identifying all conference records from Ovid Embase that would be suitable for use in systematic reviews. The method should be appropriate for both including or excluding conference records to ensure they could be reviewed comprehensively or identified separately. The objectives were to:

- understand how the *Records From* limit operates in Ovid Embase
- compare the effect of using *Records From* Embase to *Records From* MEDLINE
- assess the comprehensiveness of the *Records From* Conference Abstracts limit
- identify any additional conference records not retrieved through the *Records From* Conference Abstracts limit
- recommend a method for comprehensively identifying conference records from Ovid Embase so that they could be included or excluded
- apply the recommended method to a case study showing how it would work in practice.

### The components of Embase

All of the data was collected on March 1 2022 using the Ovid segment “Embase 1974 to 2022 February 28”. The command *docz*.*dz* was used to retrieve the entire contents of the database to test the limits and commands against the whole of Embase. The database field (.db) is a shortcut to applying the *Records From* limit that obtains identical results.

The *Records From* limit was applied to each of the categories defined in Table 1. These results were compared to show the different methods for isolating the content unique to MEDLINE in a set of search results. Each item from MEDLINE has a PubMed identity number, which can be searched in the (.pm) field of Embase. This field was used when calculating the overlap and the number of records unique to either Embase or MEDLINE.

### Identifying conference records

In Embase, a *conference abstract* is an individual presentation or poster presented at a conference, symposium or similar event and usually published in a supplement to a journal indexed by Embase.^12^ A *conference review* is a record created for Embase with details about the conference and the number of abstracts attached to it.^12^ A *conference paper* is an “original article reporting data presented at a conference or symposium”.^13(p6)^

The Ovid guide to Embase was reviewed to understand how conference records may have been indexed.^12^ The fields identified from this guide were tested to see whether they retrieved records that were not found by using the *Records From* Conference Abstracts limit. The process was swapped over to show what would be retrieved by the *Records From* limit but not the other fields. Further testing was done to understand how to exclude unique MEDLINE content to retrieve Elsevier-indexed conference records from Embase. Some manual checks of the Embase records were undertaken to verify that the results did appear to be conference abstracts, such as looking for the words colloquium, conference, congress, proceedings or symposium anywhere in the record.

The most effective combination of these methods was used in a single search line. The purpose was to create the shortest search strategy possible, which would be quicker and easier to apply.

A case study of undertaking a systematic search on COVID-19 was used to show how the recommended method could be used in practice. This was chosen as it is a current issue of international importance.

## Results

### The components of Embase

There were 36,986,580 records on Ovid Embase on March 1 2022 using the segment 1974 to 2022 February 28 (see Table 2, Line 1). This is comprised of the four document collections: using *Records From* Conference Abstracts has 4,348,438 results (Table 2 Line 2), *Records From* Embase has 23,889,111 results (Table 2, Line 4), *Records From* MEDLINE has 8,739,490 results (Table 2 Line 6), and *Records From* Preprints has 9226 results (Table 2, Line 8). The database field (.db) retrieves the same results as the *Records From* limit (Table 2, Lines 2-9). There is no overlap between any of the four components (Table 2, Lines 10-15).

**Table 2.** Number of results from the *Records From* limit in Ovid Embase 1974 to 2022 February 28

| # | Searches | Results |
| --- | --- | --- |
| 1 | docz.dz. | 36986580 |
| 2 | limit 1 to conference abstracts | 4348438 |
| 3 | conference abstracts.db. | 4348438 |
| 4 | limit 1 to embase | 23889111 |
| 5 | embase.db. | 23889111 |
| 6 | limit 1 to medline | 8739490 |
| 7 | medline.db. | 8739490 |
| 8 | limit 1 to "preprints (unpublished, non-peer reviewed)" | 9226 |
| 9 | preprint.db. | 9226 |
| 10 | 2 and 4 | 0 |
| 11 | 2 and 6 | 0 |
| 12 | 2 and 8 | 0 |
| 13 | 4 and 6 | 0 |
| 14 | 4 and 8 | 0 |
| 15 | 7 and 9 | 0 |
| 16 | 3 or 5 or 7 or 9 | 36986265 |
| 17 | 1 not 16 | 315 |
| 18 | (1* or 2* or 3* or 4* or 5* or 6* or 7* or 8* or 9* or 0*).pm. | 25963843 |
| 19 | 2 and 18 | 0 |
| 20 | 4 and 18 | 17224353 |
| 21 | 6 and 18 | 8739490 |
| 22 | 8 and 18 | 0 |
| 23 | 4 not 18 | 6664758 |

A small proportion of records are not contained in any of the four document collections. There were 36,986,580 records in total and the four components contained 36,986,265 (Table 2, Line 16), a discrepancy of 315 (Table 2, Line 17).

The unique content from the MEDLINE supplement accounts for 23.63% of records available on Embase (Table 3). These are the records that have MeSH terms automatically mapped to Emtree. The *Records From* Embase limit contains 23,889,211 records (Table 2, Line 4), comprising items that can be found on MEDLINE and others unique to Embase. There are 17,224,353 records in the *Records From* Embase limit that have a PubMed ID number (Table 2, Line 20), which means, although found on both databases, they have been fully indexed by Elsevier for Embase. A further 6,664,758 records in the *Records From* Embase limit (Table 2, Line 23), comprising 18% of the total content (Table 3), do not have a PubMed ID number. These are from journals indexed by Elsevier for Embase that cannot be found on MEDLINE.

**Table 3.** Composition of Ovid Embase 1974 to 2022 February 28

| Component | Number | Percentage |
| --- | --- | --- |
| *Records From* Conference Abstracts<br>(unique to Embase) | 4,348,438 | 11.7568% |
| *Records From* Embase<br>(unique to Embase) | 6,664,758 | 18.0194% |
| *Records From* Embase<br>(overlapping with MEDLINE and<br>indexed separately by both databases) | 17,224,353 | 46.5692% |
| *Records From* MEDLINE<br>(unique to MEDLINE) | 8,739,490 | 23.6288% |
| *Records from* Preprints<br>(unique to Embase) | 9226 | 0.0249% |
| Records unassigned to a document<br>collection | 315 | 0.0009% |
| Total | 36,986,580 | 100% |

The *Records From* Conference Abstracts limit accounts for 11.76% of the total Embase records (Table 3). Preprints account for 0.0249% of Embase (Table 3) but only two months’ worth of records had been entered at the time of testing and this is likely to increase.

Using the *Records From* MEDLINE limit to remove the unique content would reduce the total records available on Embase from 36,986,580 to 28,247,090 (Table 4, Line 5). On the other hand, applying the *Records From* Embase limit reduces the total records available to 23,889,111 (Table 4, Line 2). There is a discrepancy of 4,357,979 between these two methods (Table 4, Line 8). This is largely due to the first method retaining the 4,357,664 records from the combination of *Records From* Conference Abstracts and *Records From* Preprints (Table 4, Lines 9-10). There is a further discrepancy of 315 (Table 4, Line 11), which is the same set of records that have not been added to any of the four document collections (Table 4, Lines 11-14).

**Table 4.** Applying the *Records From* MEDLINE limit in Ovid Embase 1974 to 2022 February 28

| # | Searches | Results |
| --- | --- | --- |
| 1 | docz.dz. | 36986580 |
| 2 | limit 1 to embase | 23889111 |
| 3 | limit 1 to conference abstracts | 4348438 |
| 4 | limit 1 to "preprints (unpublished, non-peer reviewed)" | 9226 |
| 5 | 2 or 3 or 4 | 28246775 |
| 6 | limit 1 to medline | 8739490 |
| 7 | 1 not 6 | 28247090 |
| 8 | 7 not 2 | 4357979 |
| 9 | 3 or 4 | 4357664 |
| 10 | 8 and 9 | 4357664 |
| 11 | 8 not 10 | 315 |
| 12 | 2 or 3 or 4 or 6 | 36986265 |
| 13 | 1 not 12 | 315 |
| 14 | 11 and 13 | 315 |

### Identifying conference records

The *Records From* Conference Abstracts limit contains the publications types (.pt) *conference abstract* (an individual presentation) and *conference review* (a record created by Elsevier about the conference). For example, the 22^nd^ European Congress on Gynaecological Oncology can be identified in *the conference name* (.nc) field with 653 records attached to it (Table 5, Line 5). These results include one record about the conference, giving its name, dates, number of papers and an overview of the proceedings (Table 5, Line 6). There are 652 records with details of the individual conference abstracts (Table 5, Line 7). All 653 records are retrieved by the *Records From* Conference Abstracts limit (Table 5, Line 8).

**Table 5.** The contents of the *Records From* Conference Abstracts limit in Ovid Embase 1974 to 2022 February 28

| # | Searches | Results |
| --- | --- | --- |
| 1 | docz.dz. | 36986580 |
| 2 | limit 1 to conference abstracts | 4348438 |
| 3 | conference abstract.pt. | 4332471 |
| 4 | conference review.pt. | 13748 |
| 5 | 22nd European Congress on Gynaecological Oncology.nc. | 653 |
| 6 | 4 and 5 | 1 |
| 7 | 3 and 5 | 652 |
| 8 | 6 or 7 | 653 |
| 9 | 2 and 5 | 653 |

The *Records From* Conference Abstracts limit does not only contain the publications types (.pt) *conference abstract* and *conference review*. The limit contains 4,348,438 records (Table 6, Line 2) and 2389 of these are neither a *conference abstract* nor a *conference review* (Table 6, Line 6). Most of these records are recent, with 2276 of the 2389 having a publication year of 2021 or later (Table 6, Line 7). The publication type *article* accounts for 2370 of the 2389 (Table 6, Line 9) and a further two are classed as *editorial* (Table 6, Line 11). The remaining 17 records have the publication type *conference paper* (Table 6, Line 13). Manually checking the 17 records suggests that they would be relevant when searching for conference records.

**Table 6.**
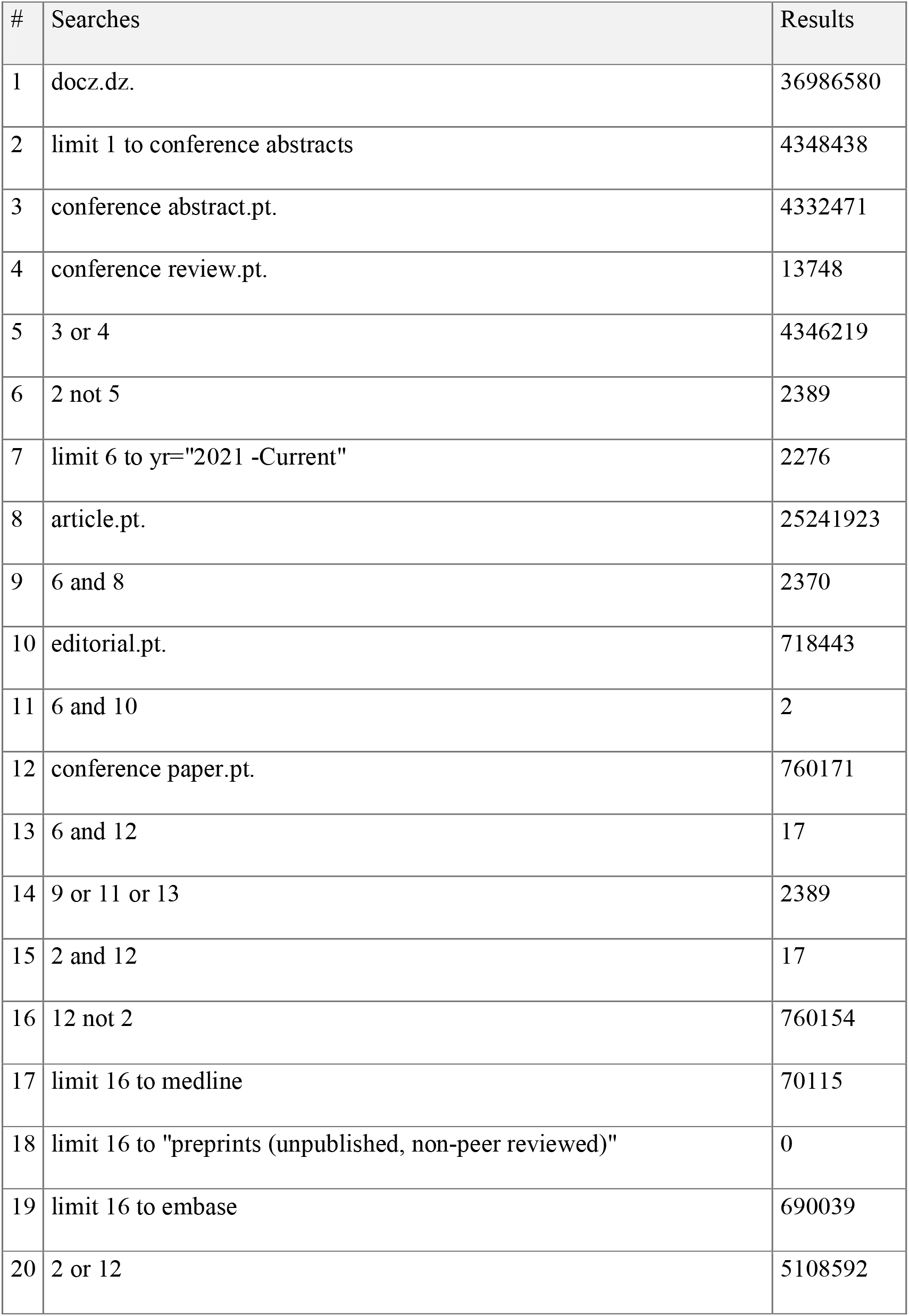
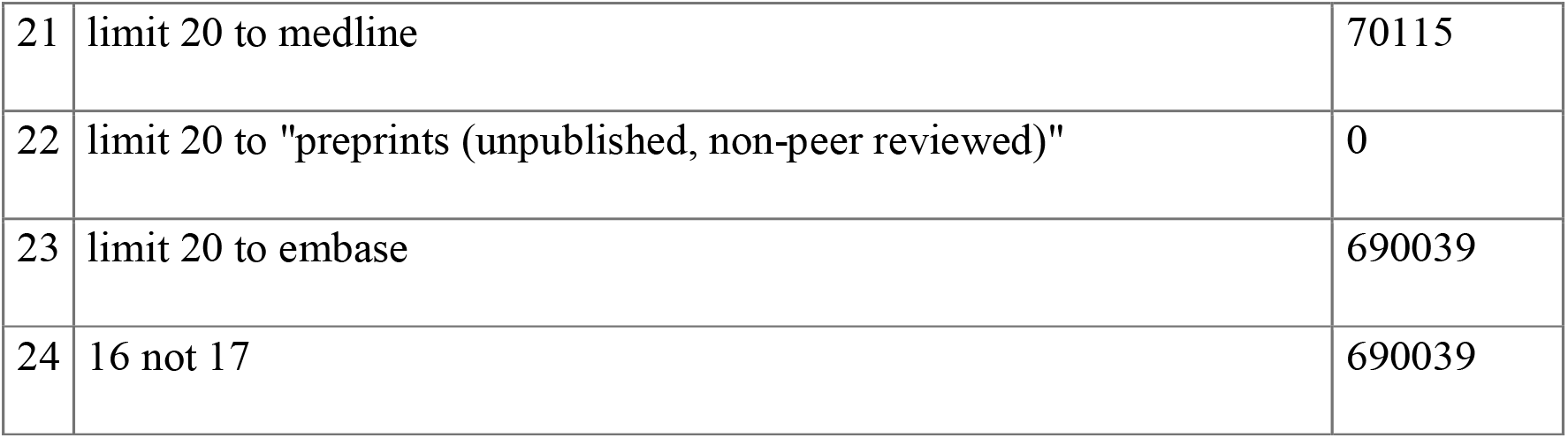
Records retrieved by the *Records From* Conference Abstracts limit and not by the *conference abstract* or *conference review* publication types in Ovid Embase 1974 to 2022 February 28

| # | Searches | Results |
| --- | --- | --- |
| 1 | docz.dz. | 36986580 |
| 2 | limit 1 to conference abstracts | 4348438 |
| 3 | conference abstract.pt. | 4332471 |
| 4 | conference review.pt. | 13748 |
| 5 | 3 or 4 | 4346219 |
| 6 | 2 not 5 | 2389 |
| 7 | limit 6 to yr="2021 -Current" | 2276 |
| 8 | article.pt. | 25241923 |
| 9 | 6 and 8 | 2370 |
| 10 | editorial.pt. | 718443 |
| 11 | 6 and 10 | 2 |
| 12 | conference paper.pt. | 760171 |
| 13 | 6 and 12 | 17 |
| 14 | 9 or 11 or 13 | 2389 |
| 15 | 2 and 12 | 17 |
| 16 | 12 not 2 | 760154 |
| 17 | limit 16 to medline | 70115 |
| 18 | limit 16 to "preprints (unpublished, non-peer reviewed)" | 0 |
| 19 | limit 16 to embase | 690039 |
| 20 | 2 or 12 | 5108592 |
| 21 | limit 20 to medline | 70115 |
| 22 | limit 20 to "preprints (unpublished, non-peer reviewed)" | 0 |
| 23 | limit 20 to embase | 690039 |
| 24 | 16 not 17 | 690039 |

The publication type *conference paper* has been applied to 760,171 records (Table 6, Line 12), of which 690,039 are in the *Records From* Embase document collection (Table 6, Line 19) and 70,115 are in *Records From* MEDLINE (Table 6, Line 17). None are categorised as preprints (Table 6, Line 18). This means a search using the *Records From* Conference Abstracts limit would only find 17 of the 760,171 records with the publication type *conference paper*. Combining these two methods gives a total of 5,108,592 conference records on Embase (Table 6, Line 20) and the *Records From* Conference Abstracts limit finds 4,348,438 (85%) of them.

There is a further set of 170 records with the publication types (.pt) *conference abstract* and *conference review* that are not contained in the *Records From* Conference Abstracts limit (Table 7, Line 6). The issue identified in Table 4 that 315 records are not included in any of the four Embase document collections accounts for 136 of the 170 missing conference records (Table 7, Line 12). Manually checking the remaining 34 records (Table 7, Line 13) suggests that they do relate to conferences.

**Table 7.** Records retrieved by the *conference abstract* or *conference review* publication types and not by the *Records From* Conference Abstracts limit in Ovid Embase 1974 to 2022 February 28

| # | Searches | Results |
| --- | --- | --- |
| 1 | docz.dz. | 36986580 |
| 2 | limit 1 to conference abstracts | 4348438 |
| 3 | conference abstract.pt. | 4332471 |
| 4 | conference review.pt. | 13748 |
| 5 | 3 or 4 | 4346219 |
| 6 | 5 not 2 | 170 |
| 7 | limit 1 to medline | 8739490 |
| 8 | limit 1 to embase | 23889111 |
| 9 | limit 1 to "preprints (unpublished, non-peer reviewed)" | 9226 |
| 10 | 2 or 7 or 8 or 9 | 36986265 |
| 11 | 1 not 10 | 315 |
| 12 | 6 and 11 | 136 |
| 13 | 6 not 11 | 34 |

The Ovid guide to Embase shows that *conference proceeding* can be used in the source type (.su) field.^12^ There are 4282 records with this source type (Table 8, Line 7), none of which overlap with the *Records From* Conference Abstracts limit (Table 8, Line 8) or the *conference abstract* or *conference review* publication types (Table 8, Lines 9-10). There is a large overlap of 4099 between source type *conference proceeding* and publication type *conference paper* (Table 8, Line 11). This leaves 183 records still to be retrieved (Table 8, Line 13), of which 34 are unique to MEDLINE (Table 8, Line 14) and 149 are from Embase (Table 8, Line 15). It is worth noting that 147 of the 149 Embase records were published before 2000 (Table 8, Line 21).

**Table 8.** Records retrieved by the *conference proceeding* source type in Ovid Embase 1974 to 2022 February 28

| # | Searches | Results |
| --- | --- | --- |
| 1 | docz.dz. | 36986580 |
| 2 | limit 1 to conference abstracts | 4348438 |
| 3 | conference abstract.pt. | 4332471 |
| 4 | conference review.pt. | 13748 |
| 5 | conference paper.pt. | 760171 |
| 6 | or/2-5 | 5108762 |
| 7 | conference proceeding.su. | 4282 |
| 8 | 2 and 7 | 0 |
| 9 | 3 and 7 | 0 |
| 10 | 4 and 7 | 0 |
| 11 | 5 and 7 | 4099 |
| 12 | 7 not 11 | 183 |
| 13 | 7 not 6 | 183 |
| 14 | limit 13 to medline | 34 |
| 15 | limit 13 to embase | 149 |
| 16 | limit 13 to "preprints (unpublished, non-peer reviewed)" | 0 |
| 17 | limit 7 to yr="1800 – 1999" | 2725 |
| 18 | limit 7 to yr="2000-2009" | 411 |
| 19 | limit 7 to yr="2010-2019" | 1146 |
| 20 | limit 7 to yr="2020-2022" | 0 |
| 21 | limit 15 to yr="1800 – 1999" | 147 |
| 22 | limit 15 to yr="2000-2009" | 0 |
| 23 | limit 15 to yr="2010-2019" | 2 |

Combining the *Records From* Conference Abstracts limit with the publication types (.pt) *conference abstract, conference review* and *conference paper* and source type *conference proceeding* has 5,108,945 results (Table 9, Line 7). This includes 70,149 records that are unique to MEDLINE (Table 9, Line 8). Removing the unique MEDLINE content means that there are 5,038,796 records relating to conferences that have been indexed by Elsevier for Embase (Table 9, Line 13), which is 690,358 higher than using the *Records From* Conference Abstracts limit alone (Table 9, Line 14).

**Table 9.** Maximizing retrieval of conference records from Ovid Embase 1974 to 2022 February 28

| # | Searches | Results |
| --- | --- | --- |
| 1 | docz.dz. | 36986580 |
| 2 | limit 1 to conference abstracts | 4348438 |
| 3 | conference abstract.pt. | 4332471 |
| 4 | conference review.pt. | 13748 |
| 5 | conference paper.pt. | 760171 |
| 6 | conference proceeding.su. | 4282 |
| 7 | or/2-6 | 5108945 |
| 8 | limit 7 to medline | 70149 |
| 9 | 3 and 8 | 0 |
| 10 | 4 and 8 | 0 |
| 11 | 5 and 8 | 70115 |
| 12 | 6 and 8 | 1555 |
| 13 | 7 not 8 | 5038796 |
| 14 | 13 not 2 | 690358 |
| 15 | (conference abstract* or conference review or conference paper or<br>conference proceeding).db,pt,su. | 5108945 |
| 16 | 7 and 15 | 5108945 |
| 17 | limit 16 to medline | 70149 |
| 18 | 16 not 17 | 5038796 |
| 19 | (conference abstract* or conference review or conference paper or<br>conference proceeding).db,pt,su. not medline.db. | 5038796 |
| 20 | limit 14 to dc=15000101-20081231 | 594427 |
| 21 | limit 14 to dc=20220101-20220228 | 938 |
| 22 | limit 2 to dc=20220101-20220228 | 46147 |
| 23 | limit 19 to dc=20220101-20220228 | 47032 |

Including the date created field (.dc) shows that 594,427 (86%) of the extra 690,358 conference records were added to Embase before 2009 (Table 9, Line 20), when Elsevier created the *Records From* Conference Abstracts limit. There is an ongoing issue, as 938 conference records added in January and February 2022 are not identified by the *Records From* Conference Abstracts limit. These represent 1.99% of the 47,032 conference records indexed by Elsevier for Embase in those two months (Table 9, Line 23).

The *Records From* Conference Abstracts limit, the publication types (.pt) *conference abstract, conference review* and *conference paper* and the source type *conference proceeding* can all be combined into a single line to make them easier to run. The *Records From* Conference Abstracts limit has to be converted to the database field (.db), which was shown in Table 2 to produce identical results. Truncation is added to account for the discrepancy between *conference abstracts* in the database field and *conference abstract* in the publication type field. Combining the searches into a single line retrieves the same 5,108,945 results as using them separately (Table 9, Lines 15-16). It is also possible to remove the unique MEDLINE content using a single line search to retrieve the same 5,038,796 records indexed by Elsevier for Embase (Table 9, Lines 18-19).

### COVID-19 case study

The National Institute for Health and Care Excellence (NICE) COVID-19 search strategy (version 7)^17^ finds 254,201 records on Embase (Table 10, Line 6), of which 32,301 are in the *Records From* Conference Abstracts limit (Table 10, Line 7). The COVID-19 set includes two records that are not part of any of the four document collections of Embase (Table 10, Line 12). Combining the *Records From* Conference Abstracts limit, the publication types (.pt) *conference abstract, conference review* and *conference paper* and the source type *conference proceeding* means that 32,731 records are retrieved (Table 10, Line 19). Records unique to MEDLINE account for 27 of the 32,731 results (Table 10, Line 20). There are 32,704 conference records indexed by Elsevier for Embase relating to COVID-19 (Table 10, Line 22).

**Table 10.**
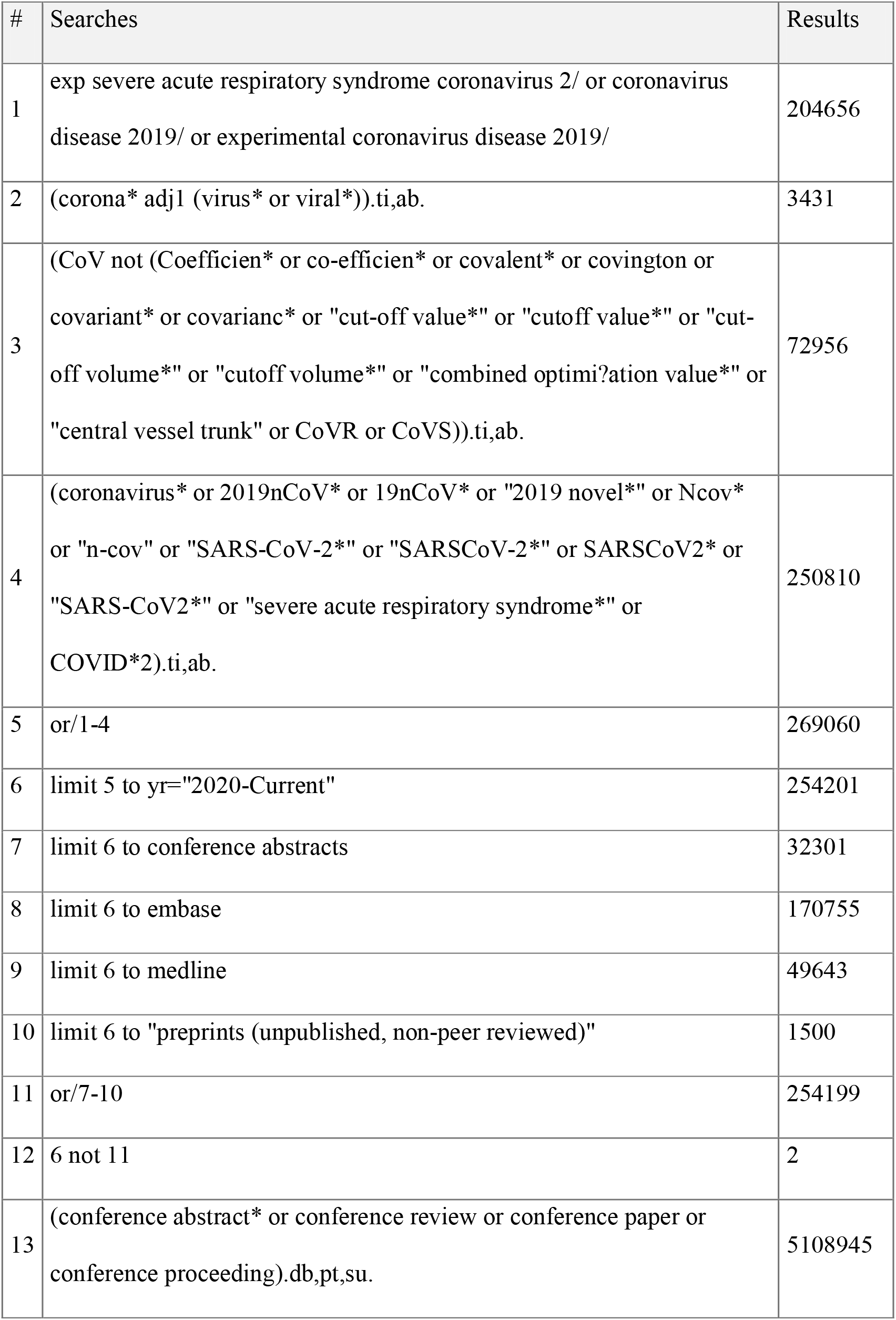
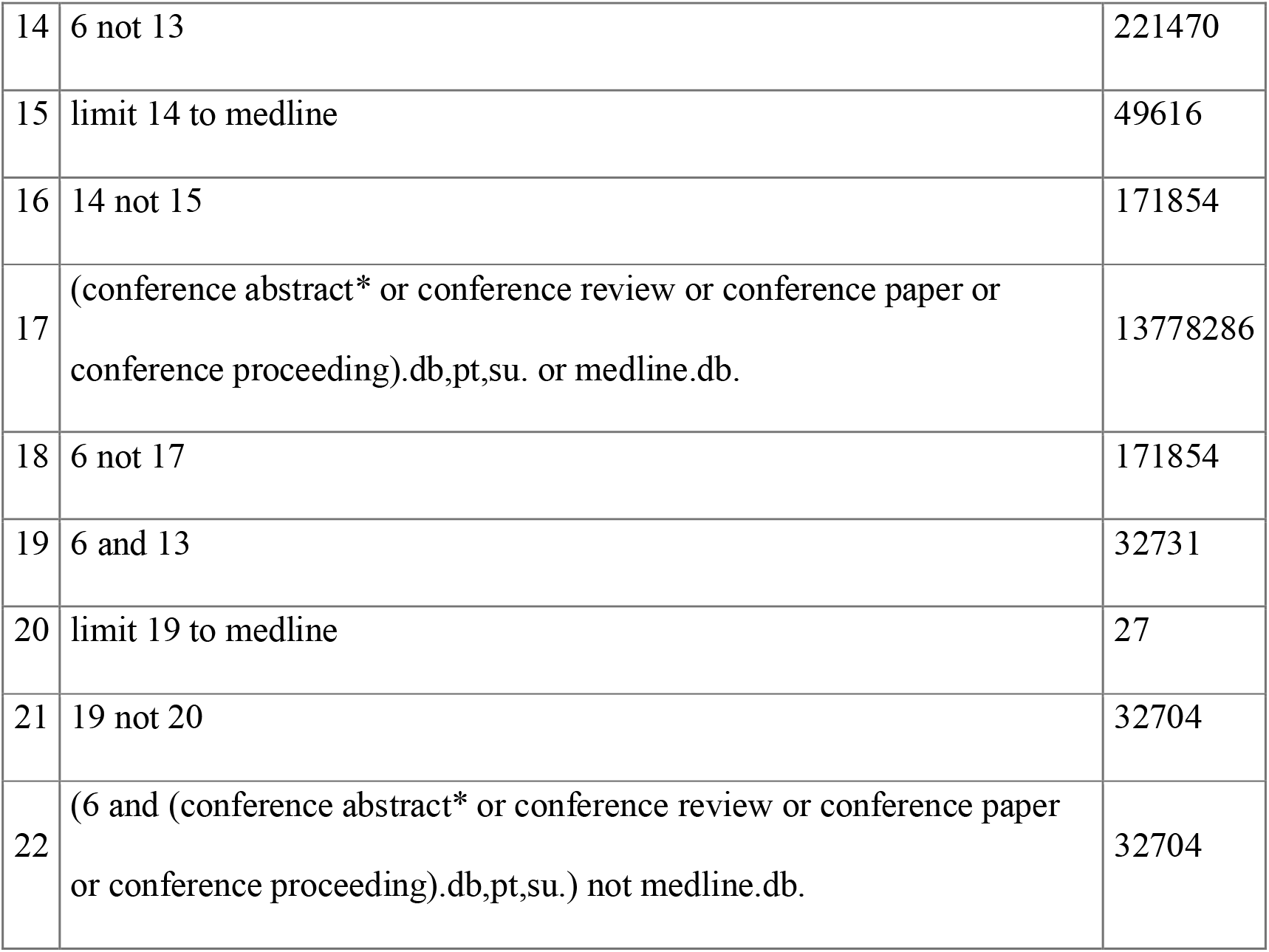
Retrieving conference records relating to COVID-19 from Ovid Embase 1974 to 2022 February 28

## Discussion

### The components of Embase

It is important to remember the definitions from Table 1 when deciding which components of Embase to use. The *Records From* MEDLINE limit includes items that are unique to MEDLINE and have not been indexed by Elsevier for Embase, although the MeSH terms have been mapped to Emtree. There is also a large set of records that overlaps between MEDLINE and Embase, as they have been indexed separately for both databases. Using *Records From* MEDLINE isolates the unique content, it does not retrieve all the content available on Embase that could also be found on MEDLINE if it were searched independently. Applying the *Records From* MEDLINE limit will reduce the number of duplicates from searching MEDLINE and Embase separately but it will not eliminate them.

Removing the unique content in the *Records From* MEDLINE limit does not obtain the same results as applying *Records From* Embase to a search. Expressed in Boolean logic, using “Search results NOT *Records From* MEDLINE” is not equivalent to using “Search results AND *Records From* Embase”. Using “Search results AND *Records From* Embase” would risk missing the unique Elsevier content contained in *Records From* Conference Abstracts, *Records From* Preprints and those records that have not been assigned to one of the document collections.

One of the reasons for searching multiple databases in a systematic review is to improve the chances of retrieving all the relevant evidence. This partly occurs because the same item will be indexed differently on each database and any limitations to the search strategy in one source could be compensated in another source.^18^ Searching Embase for a systematic review that also uses MEDLINE separately has two purposes:

- to identify records that are unique to Embase; and
- to identify records available on MEDLINE that are indexed by Elsevier with Emtree for Embase.

Removing the content unique to *Records From* MEDLINE excludes items that Elsevier has not fully indexed for Embase, even though the MeSH terms will have been automatically mapped to Emtree. This operation does not affect the records that Elsevier has fully indexed with Emtree that can also be found on MEDLINE, a set of over 17 million records comprising 46.57% of Embase (Table 3).

Embase is comprised of four document collections (MEDLINE, Embase, Conference Abstracts and Preprints) but a small proportion of records (0.0009% in Table 3) have not been added to any of them. This is a minor issue but systematic searches aiming to be comprehensive need to understand exactly what they are, or are not, retrieving when using the *Records From* limits. For example, it affects two records relating to COVID-19 (Table 9). Elsevier will eventually correct these records and assign them to the appropriate document collections. It is hoped that Elsevier would be able to eliminate the underlying operational issue in the future. Excluding *Records From* MEDLINE retains these records in the Embase search results.

The recommendation to use “Search results NOT *Records From* MEDLINE” rather than “Search results AND *Records From* Embase” is slightly different to the method tested in an earlier paper.^16^ Premji and Ganshorn acknowledge that using *Records From* Embase should only be used when conference records are not required or are being searched separately. Their research was also undertaken before the preprints document collection was added to Embase.

These factors highlight the continuing benefit of searching MEDLINE and Embase separately for systematic searches, even if a large proportion of MEDLINE is made available via Embase.

### Identifying conference records

The testing shows that to get complete coverage of conference records on Ovid Embase it is necessary to include:

- The *Records From* Conference Abstracts limit
- The *conference abstract* publication type
- The *conference review* publication type
- The *conference paper* publication type
- The *conference proceeding* source type.

The database field (.db) provides identical results to the *Results From* limit. This means that the five elements of the search can be combined into the following line:

> (conference abstract* or conference review or conference paper or conference proceeding).db,pt,su.

This line can be used to either include or exclude conference records from a set of search results. For example, in a scenario where it is helpful to process the results separately, this line could be applied with AND to download the conference records and then it could be applied with NOT to access the other, non-conference records.

It is not strictly necessary to combine all of the search terms with all of the search fields. *Conference review, conference paper* and *conference proceeding* are not indexed in the database (.db) field. *Conference abstract, conference review* and *conference paper* are not indexed in the source type (.su) field. *Conference proceeding* is not indexed in the database (.db) or publication type (pt) fields. This does not affect the search results and it is less cumbersome than using each term separately.

The majority (86%) of the additional conference records identified by the recommended method were added to Embase before the *Records From* Conference Abstracts limit was created in 2009. The recommended method is not, however, just beneficial when searching for pre-2009 conference records. Using the *Records From* Conference Abstracts limit would miss 1.99% of the conference records indexed by Elsevier and added to Embase in the first two months of 2022 (Table 9). The publication type *conference paper* was in use before 2009 and it can still be applied to items that are neither *conference abstracts* nor *conference reviews* (the two inclusion criteria for the *Records From* Conference Abstract limit).^13^

Some of the terms in the recommended method make minimal impact on the results. The source type *conference proceeding* adds 183 conference records (Table 8, Line 13), to take the total from 5,108,762 (Table 8, Line 6) to 5,108,945 (Table 9, Line 6). Including this term in the search only adds two records published since 2000 to the conference records indexed by Elsevier for Embase. All of the possible search terms have been included, as the purpose was to identify conference records comprehensively.

It is possible (but optional) to identify the conference records from the unique MEDLINE content in a single operation, using the following strategy:

> (conference abstract* or conference review or conference paper or conference proceeding).db,pt,su. and medline.db

The recommended methods for retrieving conference records from Ovid Embase are set out in Table 11. The table shows the scenarios that are most likely to be encountered in systematic searches. Searches for conference records can be done with or without the Embase content that is unique to MEDLINE. There is a long-form method, where it is important to observe the effects separately, and a short-form method where the intermediate steps are not recorded. Further work would be required to test these methods in the versions of Embase hosted on other platforms.

**Table 11.**
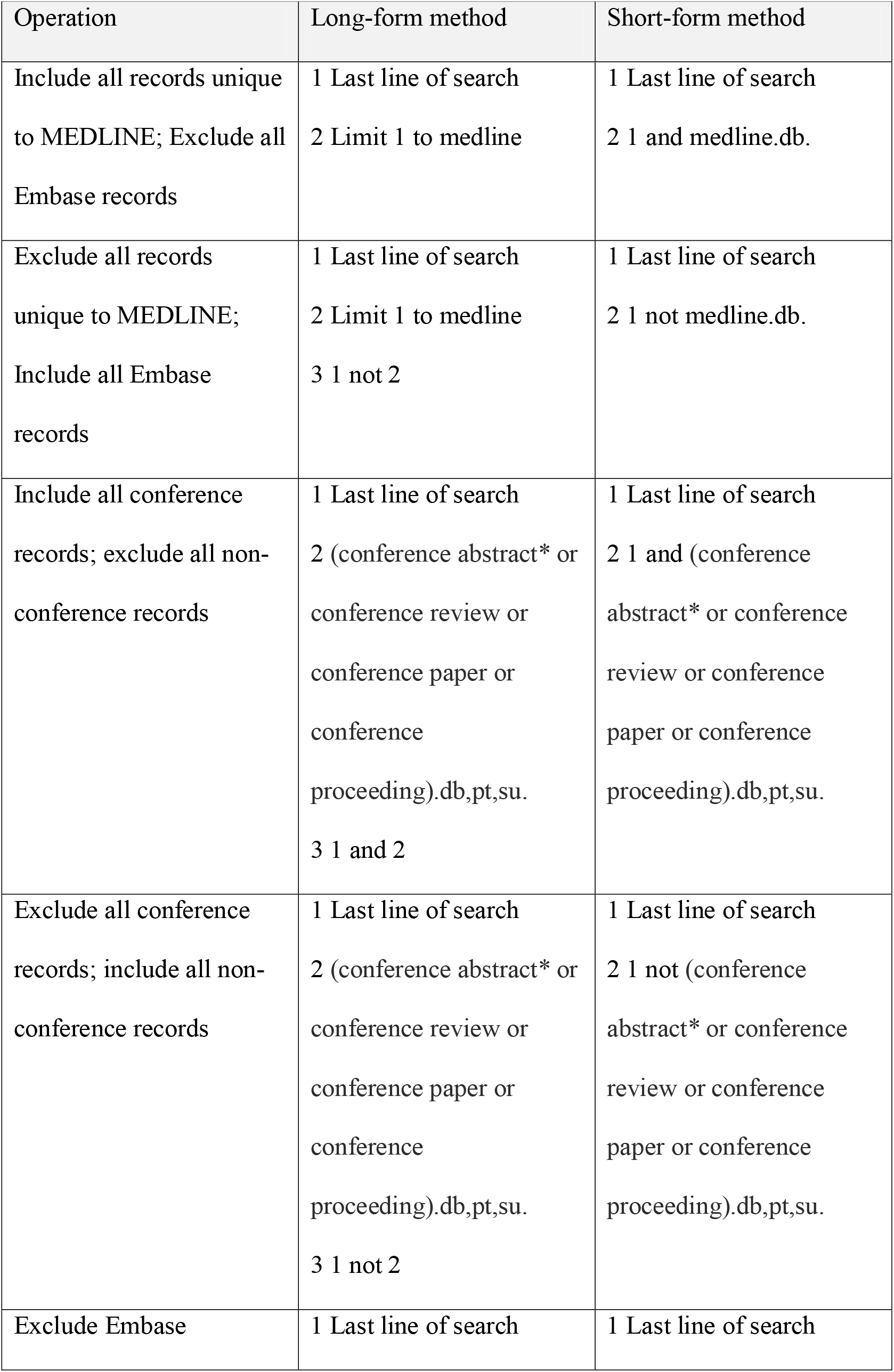
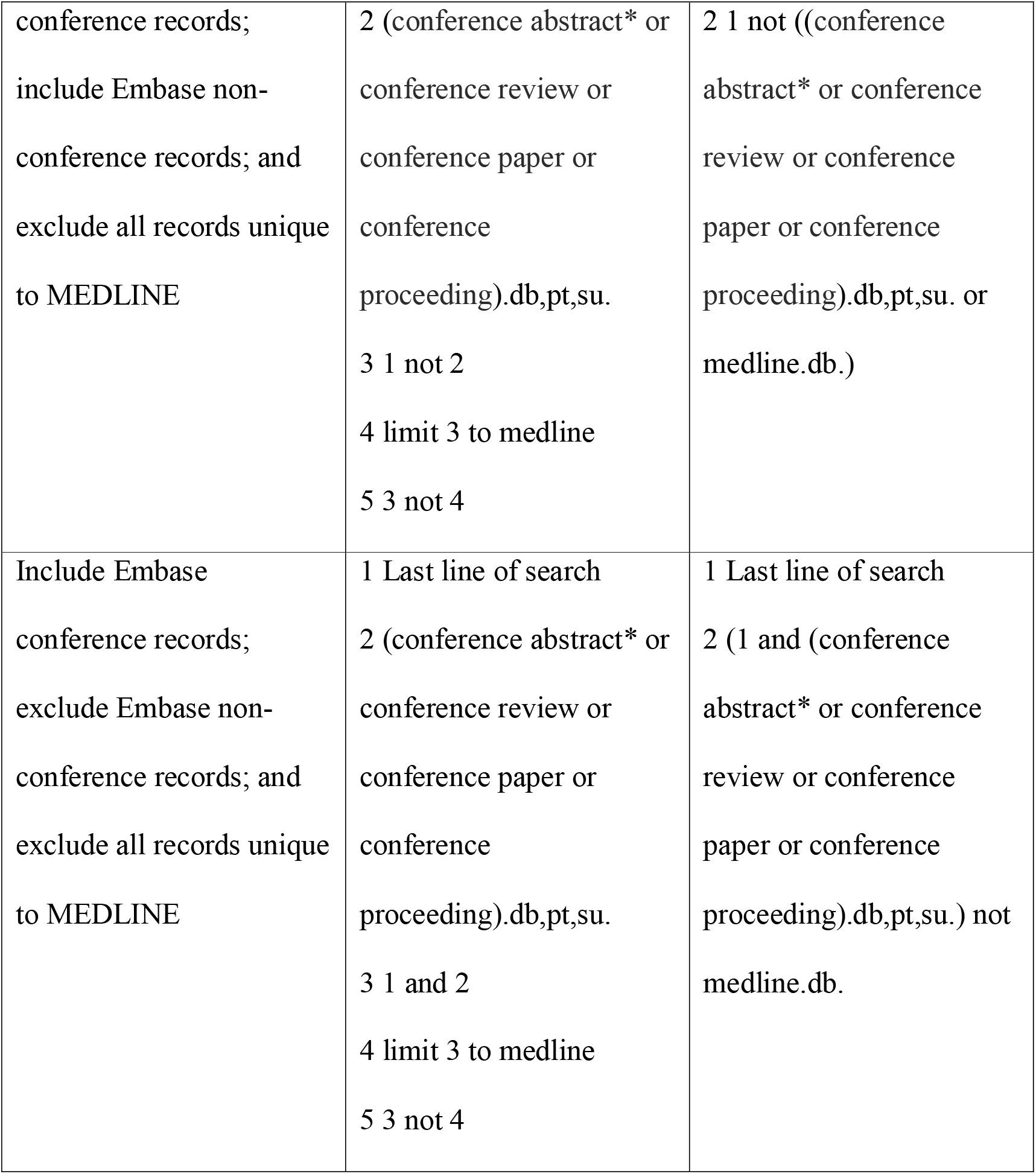
Recommended methods for using the *Records From* limit and identifying conference records in Ovid Embase

## Conclusions

The testing, undertaken on March 1 2022, agrees with Premji and Ganshorn that the *Records From* Embase limit is only suitable if conference records are not required in the search results.^16^ This paper has found that it is more reliable to use the format “Search results NOT *Records From* MEDLINE”, as this will retain conference records, preprints and items not yet added to a document collection.

The testing found 5,108,945 conference records on Embase, from a total of 36,986,580 records (around 14%). The recommended method combines the *Records From* Conference Abstracts limit, the publication types (.pt) *conference abstract, conference review* and *conference paper* and the source type *conference proceeding*. Once the unique MEDLINE content was excluded, the recommended method finds 5,038,796 conference records that had been indexed by Elsevier for Embase, which is 690,358 higher than using the *Records From* Conference Abstracts limit alone. The figures will obviously fluctuate as more records are added on a daily basis but they show that Embase is a significant source of conference records.

## Data Availability

All the data that supports the findings of this study are available in the manuscript.

## Acknowledgements

The author would like to thank Amy Finnegan, Sarah Glover, Tom Hudson, Caroline Miller, Nicola Walsh and Ceri Williams for their assistance. Thanks also to the Ovid technical support team for answering various queries that have helped to inform the development of this paper. The author is grateful to Pieter van der Houwen, Product Strategy Manager at Wolters Kluwer, for commenting on an earlier draft of this paper.

## Availability of data

All the data that supports the findings of this study are available in the article.

